# Body surface gastric mapping parameters are associated with response to gastric peroral endoscopic myotomy for gastroparesis: pilot study

**DOI:** 10.1101/2025.09.18.25336055

**Authors:** Homira Ayubi, Chris Varghese, Mabel Tanne, Gabriel Schamberg, Shraddha Gulati, Mehul Patel, Amyn Haji, Greg O’Grady, Bu’Hussain Hayee

**Affiliations:** King’s College Hospital, London, United Kingdom; University of Auckland, Auckland, New Zealand; Mayo Clinic, Rochester, Minnesota, United States of America; Alimetry Ltd, Auckland, New Zealand

## Abstract

**Message:** Gastric per oral endoscopic myotomy (GPOEM) is a promising therapy for refractory gastroparesis, but patient selection remains challenging. We evaluated body surface gastric mapping (BSGM) phenotypes to predict treatment response. Patients were recruited at King’s College Hospital (Nov 2022–July 2025). BSGM comprised 30-min fasting, standardized nutrient drink with oatmeal bar (482 kcal), and 4-h postprandial recording. Success was defined as ≥1 point reduction in Gastroparesis Cardinal Symptom Index or complete symptom resolution at follow-up. Overall, 53% responded, including all patients with dysrhythmic or continuous phenotypes. Higher gastric frequencies predicted non-response (p=0.03).

## Background

Gastroparesis is a heterogeneous condition, often associated with a markedly reduced quality of life yet with few effective therapies. Gastric per oral endoscopic myotomy (GPOEM) has emerged as a promising treatment for refractory gastroparesis,^1^ but patient selection remains challenging, with response rates rarely exceeding ∼60%.^2^ Multiple mechanisms contribute to gastroparesis pathophysiology,^3,4^ and deep phenotyping of underlying pathophysiologies may augment patient selection for GPOEM. We therefore assessed the ability of body surface gastric mapping (BSGM) using Gastric Alimetry® (Auckland, New Zealand), a non-invasive test of gastric function, to predict symptomatic response to GPOEM.

## Methods

### Patient eligibility

Patients with suspected gastroparesis at Kings College Hospital were recruited (Nov 2022 -July 2025).

### Outcomes

Success was defined as either a 1-point reduction in gastroparesis cardinal symptom index (GCSI) or complete resolution of symptoms. GCSI measurements were performed with follow-up, and data closest to 6-months were used when multiple time-points were available.

### Device and technique

Gastric Alimetry (Auckland, New Zealand) was used for BSGM and symptom capture prior to GPOEM. BSGM included 30 m fasting, standard nutrient drink + oatmeal bar (482 kcal), and 4 h postprandial recording. BSGM phenotyping followed an expert consensus incorporating BSGM spectral biomarkers, symptoms, and clinical histories (Auckland Classification v1.0; in development).

### Phenotypes included

- *Dysrhythmic phenotype*: low rhythm stability based on Gastric Alimetry Rhythm Index (GA-RI; <0.25).^5^ Associated with interstitial cell of Cajal (ICC) depletion or dysfunction.^6^
- *High frequency phenotype*: high Principal Gastric Frequency (PGF; >3.4, i.e., in outside the 95% confidence intervals of the normative reference intervals, refer ^5^). Associated with vagal neuropathy.^7,8^
- *Low meal response phenotype*: defined as a meal response ratio < 1 (i.e., ratio of BMI-adjusted amplitude in the first 2 postprandial hours to the last 2 hours during the 4 h recording).^3^ Low meal response ratio seen in association with delayed emptying,^3^ and relative postprandial hypomotility on BSGM has shown to benefit from prokinetics.^9^
- *Sensorimotor phenotype*: high correlation between BMI-adjusted amplitude and symptom curves (≥ 1 symptom; correlation coefficient >0.5).^10^ Considered to be associated with visceral hypersensitivity.^4^
- *Continuous phenotype*: high constant symptom burden unrelated to meal or gastric motor activity.^10^ Associates most strongly with psychological comorbidity in large BSGM datasets.^11^

Remaining patients were grouped as *‘Other’*.^12^

#### Data analysis

Analyses were performed in R v4.0.3 (R Core Team, Vienna, Austria). Distributions were assessed. Wilcoxin signed rank tests were used for non-normally distributed paired comparisons, t-tests were used for normally distributed comparisons. Categorical data were compared via Fischer’s exact test. A binomial logistic regression was constructed to investigate the association between spectral metrics from BSGM (BMI-adjusted amplitude, GA-RI, and PGF) to GPOEM success criteria with adjustment for age, BMI and sex. A restricted cubic spline with 3 knots was plotted to visualise the change in odds ratio across the range of gastric frequencies.

## Results

30 patients completed follow-up (median 7.1 months; 77% female; median age 38.5; IQR 30-52). Gastroparesis aetiologies included 11 (36.7%) diabetic, 18 (60%) idiopathic, and 1 post-surgical. BSGM phenotyping showed 3 (10%) with a ‘Dysrhythmic’ phenotype, 4 (13.3%) with a ‘High frequency’ phenotype, 9 (30%) with ‘Low meal response’ phenotype, 2 (6.7%) with a ‘Continuous’ phenotype, and 5 (16.7%) with a ‘Sensorimotor’ phenotype. Seven patients were classified as ‘Other’ with 1 including a low frequency secondary to foregut surgery, and 5 with normal tests (**Figure 1**).

**Figure 1:**
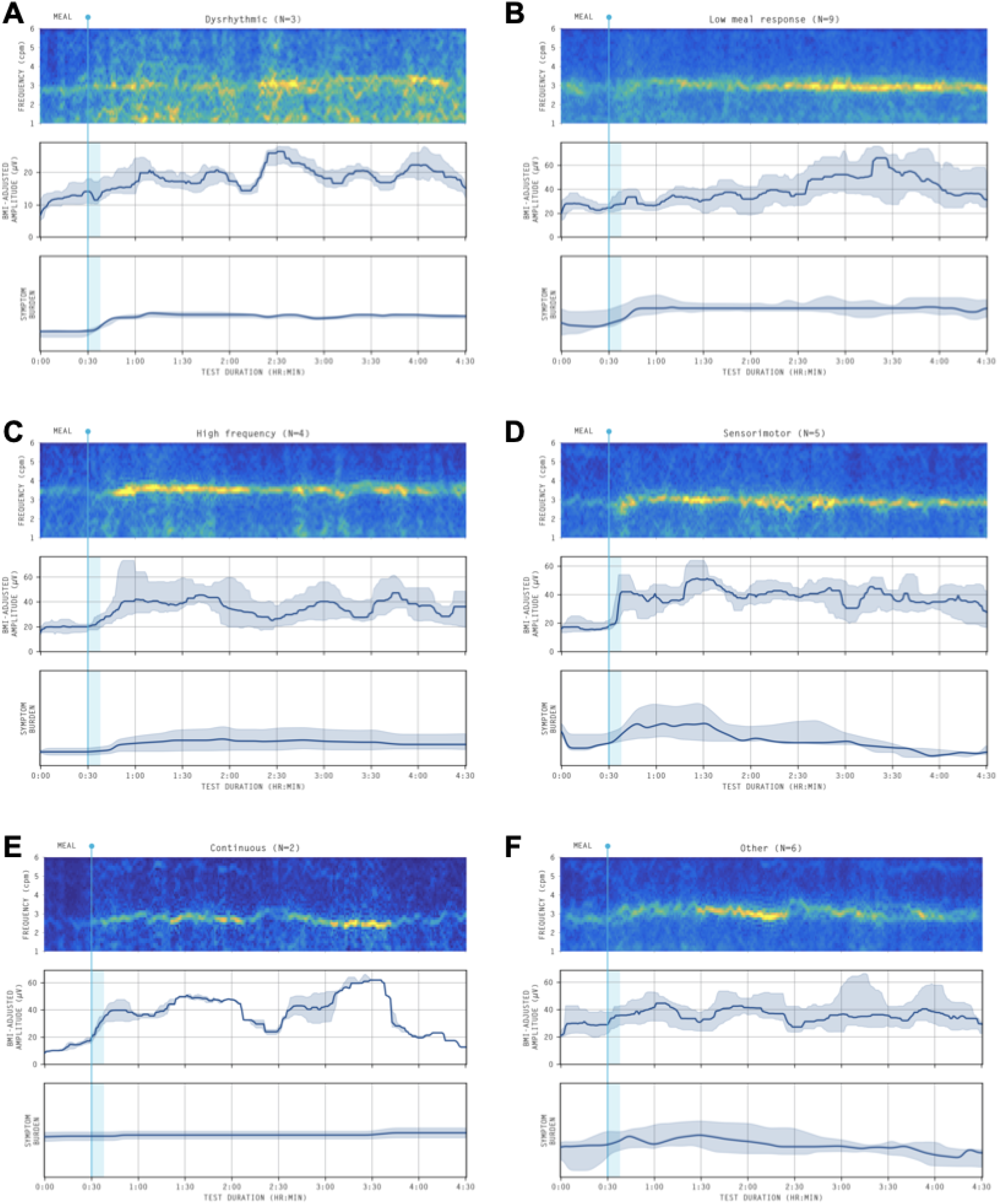
Average amplitude-frequency spectrogram, BMI-adjusted amplitude curve and average total symptom burden curve for each phenotype. A) Dysrhythmic phenotype; B) Low meal response phenotype (weak post-prandially); C) High frequency phenotype; D) Sensorimotor phenotype; E) Continuous phenotype, F) Other. Single case with low frequency phenotype secondary to foregut gastrectomy not included.

Overall, GCSI scores significantly improved following GPOEM (median -0.99, IQR -1.98 to -0.22, p<0.001; **Figure 2**), with 16 patients (53.3%) meeting the response criteria. Response rates varied significantly by phenotype (p=0.035). All patients with a ‘High frequency’ phenotype failed GPOEM. This was supported by a higher mean Principal Gastric Frequency in non-responders: 3.2 (SD 0.3) vs 2.9 (SD 0.2) in responders (p=0.02) corresponding to a reduced adjusted odds ratio (aOR) for success (aOR 0.01, 95% confidence interval 0.00-0.39, p=0.03; **Figure 3**). Meanwhile, all patients with a ‘Dysrhythmic’ or ‘Continuous’ phenotype responded (**Figure 4**).

**Figure 2:**
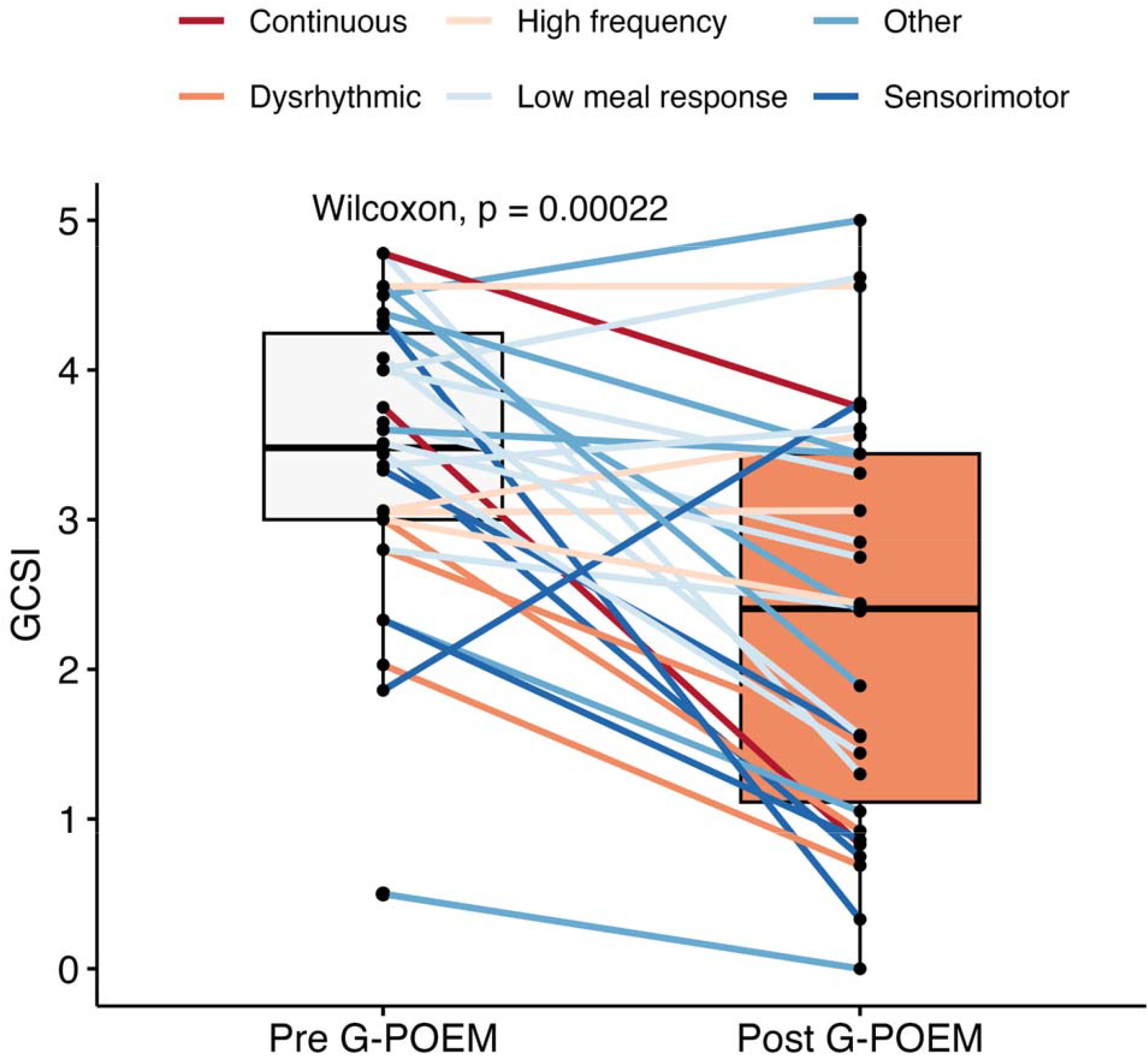
Difference in gastroparesis cardinal symptom index (GCSI) before and after gastric per oral endoscopic myotomy (*p*-value from a paired Wilcoxon signed rank test).

**Figure 3:**
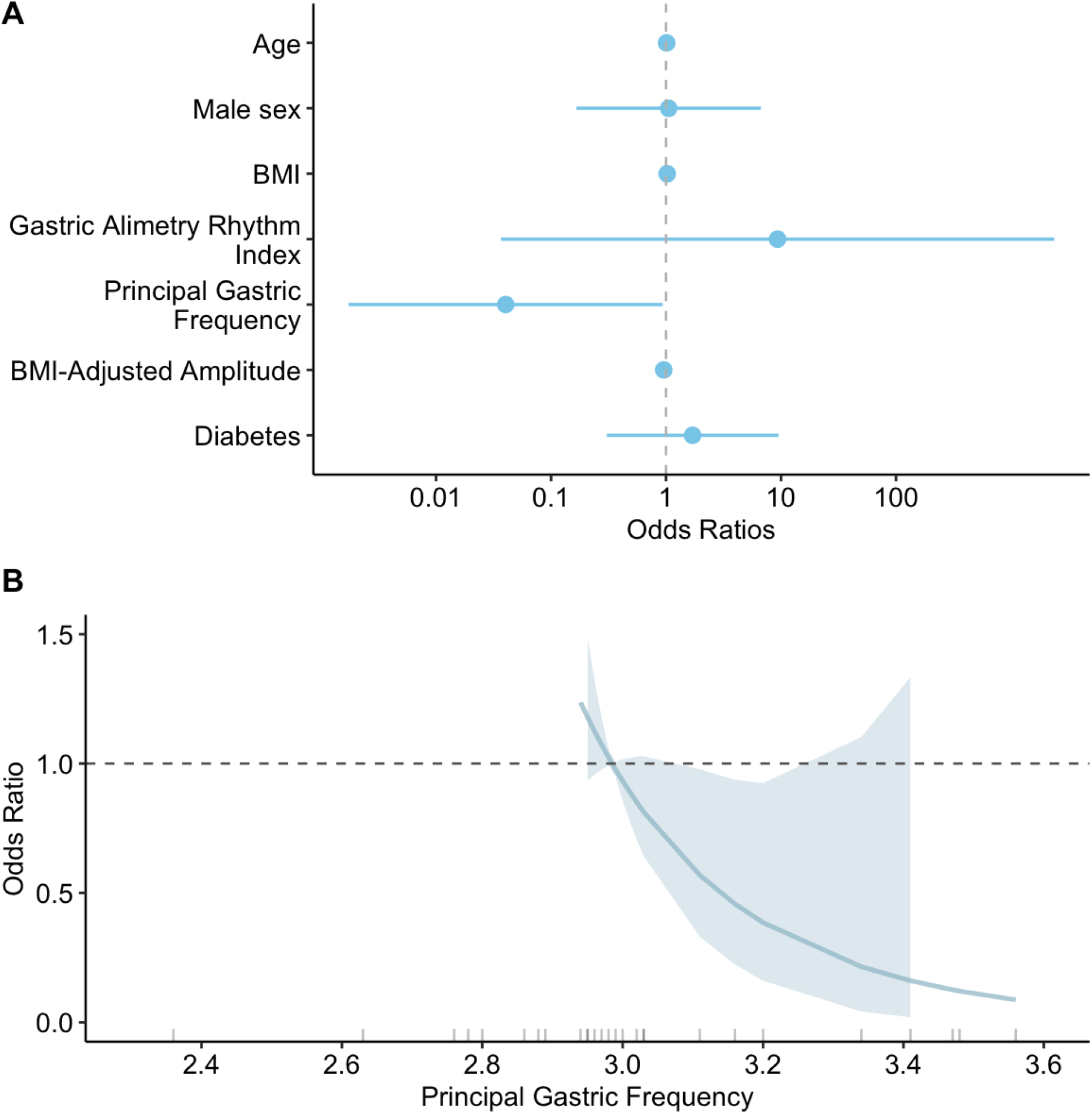
A) Forest plot for binary logistic regression model for GPOEM success including covariates age, sex, body mass index, comorbid diabetes, and each spectral metric (BMI-Adjusted Amplitude, Principal Gastric Frequency (PGF), Gastric Alimetry Rhythm Index); B) Restricted cubic spline plotting declining odds ratio as PGF increases (p=0.03).

**Figure 4:**
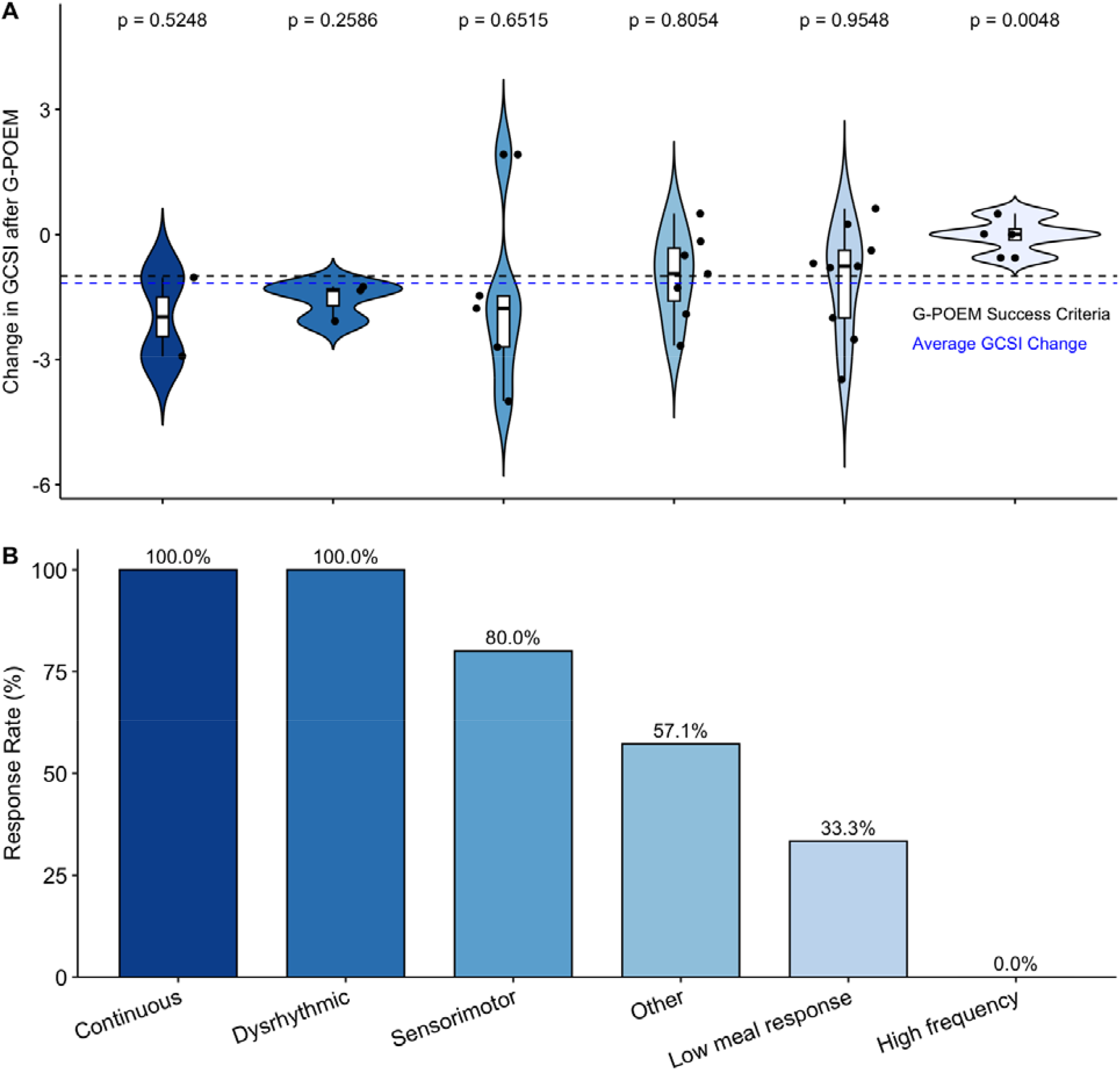
A) Violin and box and whisker plots for change in gastroparesis cardinal symptom index (GCSI) from pre to post-gastric per oral endoscopic myotomy (GPOEM). P-values above each phenotype correspond to *t*-tests comparing the average GCSI change in each phenotype to the overall group average change in GCSI; B) Bar plot of percentage response rate by phenotype (p=0.035).

## Discussion

This study demonstrates that deep phenotyping in gastroparesis using BSGM parameters offers a promising pathway to predict GPOEM outcomes. The most consistent finding was that patients with a dysrhythmic phenotype, considered to be associated with ICC-opathies, universally responded to GPOEM, exceeding the overall cohort average improvement in GCSI. By contrast, high gastric frequency, commonly observed in diabetic neuropathy,^7,10^ was associated with the poorest symptomatic response to GPOEM.

This pilot study supports a growing literature suggesting that gastroparesis care may be improved through deep phenotyping to target underlying mechanisms.^9,13^ The benefit in patients with sustained dysrhythmia is conceivably mediated through reducing pyloric resistance and intra-gastric pressure, promoting transit in the context of immune activation-driven ICC depletion.^6,14^ Positive response to GPOEM in this group is particularly advantageous given that this group currently lacks disease-modifying therapy and may fail prokinetics.^9^ Notably, those with coordinated rhythms, but low postprandial amplitudes did not benefit, and approaches employing prokinetics to augment their weak but coordinated motility could be prioritised.^9^

Patients with the high frequency phenotype, particularly at the higher ranges, are associated with treatment refractory symptoms.^7^ This phenotype is most frequently seen in association with longstanding diabetes, and is currently thought to be underpinned by vagal neuropathy.^8,15^ When seen in association with neuropathic pain and longstanding poor glycaemic control, this group is emerging as a negative predictor for pylorus-directed interventions. This was recently corroborated in a systematic review of predictors of GPOEM outcomes that found diabetes was generally associated with treatment failure.^2^

Two sensory-predominant groups also responded to GPOEM in this pilot study. The sensorimotor phenotype, in which symptoms and gastric activity are correlated, demonstrated high response rates. More studies are now needed to understand whether this association may relate to visceral hypersensitivity or dysaccommodation, both of which could potentially benefit from reduced intra-gastric pressure. Whilst patients with a continuous phenotype also responded, these results are exaggerated by a single hyperresponder and a borderline responder and therefore require further verification.

This pilot study is limited in sample size, with a median follow-up of 7 months. Hence, further validation of these findings with longer-term data is currently underway through the prospective multicentre GPOEM-GEMS study (NCT06381349), currently recruiting in >7 centres across 3 continents. Further, clinical phenotyping schemes for BSGM are currently evolving, with a consensus classification in progress.

In summary, this pilot study showed that response to GPOEM varied by BSGM phenotype, offering a novel approach to patient selection. Robust validation of the present findings is now underway.

## Data Availability

All data produced in the present study are available upon reasonable request to the corresponding author.

## Acknowledgements

N/A

